# Modelling the health and budget impact of introducing diphtheria-tetanus-pertussis-containing-vaccine booster doses in Uganda

**DOI:** 10.64898/2026.10.15.26365602

**Authors:** Rachel A. Hounsell, Sheetal P. Silal, Jared Norman, Retselisitsoe Monyake, Ampeire Immaculate, Albert Besigye, Godfrey Biroma, Joseph Magoola, Nishant Kishore, Sarah E. Pallas, Kirsten Ward

## Abstract

Routine infant vaccination has helped reduce cases and deaths from diphtheria, tetanus, and pertussis globally. However, these diseases continue to pose serious health risks as protective immunity wanes over time. Three booster doses of diphtheria-tetanus-pertussis-containing vaccine (DTPCV) administered from 12-months to 15-years of age increase duration of protection beyond childhood. Growing demands on government health spending underscore the importance of understanding the health and economic impact of vaccines prior to introduction.

Using an integrated multi-disease epidemiological-economic model, we estimated the health impact (cases averted, deaths averted, and changes to population immunity) and budget impact of introducing DTPCV booster doses in Uganda.

Compared to no DTPCV boosters, introducing all three booster doses is projected to result in 2.2 times the proportion of the population protected against symptomatic diphtheria and 2.8 times against tetanus infection among Ugandans under 15 years of age by 2039. Compared to introduction of the early childhood booster (EC) at 18 months of age, introducing all three DTPCV boosters simultaneously is projected to avert 3.2 times the number of tetanus cases over fifteen years at 2.4 times the incremental cost. The projected government vaccination cost per planned dose was USD 0.58 for simultaneous introduction of all three boosters compared with USD 0.68 for the EC booster alone. The average annual incremental cost of all three DTPCV booster doses is USD 2.79 million. Under modelled assumptions, simultaneous introduction of all three DTPCV booster doses in Uganda provides the greatest health gains, with a lower government vaccination cost per planned dose than introduction of the EC booster alone, but may still pose affordability challenges for Uganda’s government.

## 1. Introduction

Diphtheria, tetanus, and pertussis are vaccine-preventable diseases of public health importance, globally [1–4]. While significant progress has been made in reducing the burden of these diseases, they continue to pose substantial health security risks. The 2019 Global Burden of Disease (GBD) study, the latest available at the time of this analysis, estimated that globally there were 73,662 cases and 34,684 deaths from tetanus; 19.5 million cases and 117,000 deaths from pertussis; and 17,775 cases and 5,123 deaths from diphtheria [5–7]. Of tetanus and diphtheria cases reported to the World Health Organization (WHO) in 2024, 73% and 62% respectively of the global reported total were from the African Region [8]. Less than 1% of global reported pertussis cases, for which many countries in Africa do not have routine surveillance and reporting, were reported from the African Region [8].

Uganda achieved elimination status for maternal and neonatal tetanus (MNT) in 2011, though since then cases of MNT have been reported each year [8]. In 2024, Uganda reported 3,094 total tetanus cases and 686 neonatal tetanus cases [8]. The tetanus mortality rate in Uganda from 2011–2020 was approximately 50% [9]. A 2016–17 study of 24,518 non-NT cases in Uganda found 90% had no documentation of tetanus vaccination prior to the wound or tetanus symptoms, highlighting the need for vaccination as a primary prevention against tetanus [10]. Since the 1990s, pertussis cases in Uganda have been reported to the World Health Organization (WHO) in only one year—382 cases in 2021 [8]. However, the 2019 GBD study estimated that there are 200,000 to 350,000 annual cases of pertussis in Uganda, suggesting substantial underreporting of the true burden [7]. In a cross-sectional study of 449 children presenting with persistent cough at the Makerere-Mulago hospital in Kampala in 2013, a high prevalence of pertussis infection was found (15% on PCR and 20% on ELISA) [11]. The 2019 GBD study estimated that there were 397 annual cases of diphtheria in Uganda [12]. However, between 2000– 2024, there was only one year in which diphtheria cases in Uganda were reported to the WHO—49 cases in 2021 [8]. The ongoing outbreaks of diphtheria globally, catalysed by low vaccination coverage, emphasise the importance of ensuring adequate population immunity, especially in children under 15 years of age who are at highest risk of severe disease [13].

The Uganda National Expanded Programme on Immunisation (UNEPI) was launched in 1983 and included vaccination against diphtheria, tetanus, and pertussis; initially with the DT-whole cell pertussis (DTwP) formulation, then in 2002 switched to the DTwP-*Haemophilus Influenzae* type B (Hib)-Hepatitis B (HepB) vaccine, known as pentavalent (Penta) vaccine. Three doses of Penta vaccine are scheduled at 6, 10 and 14 weeks of age (the primary vaccination series). Since 2010, national coverage of the first Penta dose in Uganda among children under 12 months of age has ranged between 93% and 99% and coverage of the third Penta dose has ranged between 85% and 94% [8].

Immunity against tetanus, diphtheria, and pertussis is not lifelong, with protection waning around five to ten years following the primary vaccination series [1–3]. To extend protection against diphtheria, tetanus, and pertussis into adulthood, three booster doses of diphtheria and tetanus-containing vaccine (DTCV) are recommended, one each at 12– 23 months of age, at 4–7 years of age, and in adolescence (9–15 years of age). A booster dose of pertussis-containing vaccine is recommended for children 1–6 years of age, preferably to be given during the second year of life and in combination with DTCV. Since 2023, countries eligible for support from Gavi, the Vaccine Alliance (Gavi), which include Uganda, can apply for funding to introduce any of the three diphtheria-tetanus-pertussis-containing vaccine (DTPCV) booster doses in the national immunisation schedule. At the time of this study, there were no DTPCV booster doses—early childhood (EC), childhood (C), or adolescent boosters (A)—in the existing routine immunisation schedule in Uganda.

The Uganda National Immunisation Technical Advisory Groups (UNITAG) considered the introduction of DTPCV booster doses into the routine immunisation schedule in Uganda during the 2025 cycle of UNITAG’s New Vaccine Prioritisation Exercise. To prioritise limited financial resources for health amid expanding vaccine recommendations, evidence on the potential impact of introducing new vaccines is essential. Prior to this study, there were no integrated epidemiological-economic models to assess the impact of DTPCV booster doses, and no application of such models in a low- and middle-income country setting. To address this need, we conducted a participatory mathematical and economic modelling process to support country-level decision-making about the introduction of DTPCV booster doses. We aimed to estimate the health and budget impact of alternative strategies for introducing DTPCV booster doses in Uganda, including different combinations of booster doses, approaches to introduction, and vaccination coverage assumptions. We achieved this aim using an integrated multi-disease epidemiological-economic model developed and applied with Uganda’s national immunisation programme.

## 2. Materials and Methods

### 2.1 Ethics Statement

This modelling study used aggregate secondary data obtained from publicly available sources and programme-level inputs and did not involve recruitment of human participants or use of individually identifiable participant data. Ethical approval and informed consent were therefore not required.

### 2.2 Integrated model overview

Through a global multidisciplinary collaboration, we developed DTP Boost, an interactive web-based decision-making tool enabling users to model the health impact, budget impact, and cost-effectiveness of introducing DTPCV booster doses in a selected country under user-defined vaccination programme designs [14,15]. Multiple vaccination strategies can be designed, allowing a detailed and interactive exploration of the relative costs and benefits of introducing different numbers of DTPCV booster doses, vaccination coverage levels, introduction approaches (e.g. simultaneous, phased), vaccine formulations and vaccination delivery platforms (e.g. health facility, outreach site).

DTP Boost is powered by an integrated epidemiological-economic model. We developed deterministic age-structured compartmental models for tetanus, pertussis, and diphtheria. As these diseases have fundamentally different processes, the three models are semi-independent and linked through vaccination delivery and population characteristics (e.g. ageing, fertility, and mortality rates). The three disease models are detailed in separate publications [16–18]. In Uganda, a multi-disciplinary technical working group (TWG) was formed to collaboratively use DTP Boost, developing country-specific vaccine introduction scenarios accounting for the existing vaccination schedule, current burden of disease, health systems characteristics, costs of illness, and vaccination. Country-specific birth and death rates, capturing population growth, were sourced from the UN Population Prospects. Contact rates were incorporated into the models using an age-group and population-adjusted contact matrix adapted from Prem et al. [19].

### 2.3 Inputs and scenarios

To ensure that model inputs were appropriate to the Ugandan context, all input data were either received from, or discussed with, staff from UNEPI. Scenarios were developed to explore key questions about policy and practice under different assumptions (Table 1). We consider different combinations of DTPCV booster doses (one, two or all three) and implementation approaches (introduction in a simultaneous, phased, or expanded approach). We also explored the impact of vaccination coverage levels and time it takes to achieve this. Further details on inputs, assumptions, and scenarios are provided in the supplementary materials.

**Table 1.** Summary of scenarios to inform key policy and practice questions for DTPCV booster dose introduction in Uganda.

| Scenario | Scenario abbreviation | Scenario Description |
| --- | --- | --- |
| Baseline | Baseline | Continuation of existing schedule (primary series and maternal vaccination only) at average of previous 3-year coverage levels with no introduction of new booster doses |

| Which DTPCV booster doses to introduce? |  |  |
| --- | --- | --- |
| Introduction is simultaneous (same year of introduction) where more than one dose is introduced |  |  |
| One dose only | EC only | Early childhood (Pentavalent at 18 months) |
| Two doses (C) | EC+C | Early childhood (Pentavalent at 18 months) + Childhood (Td at 6 years) |
| Two doses (A) | EC+A | Early childhood (Pentavalent at 18 months) + Adolescent (Td at 10 years) |
| Three doses^ | EC+C+A | Early childhood (Pentavalent at 18 months) + Childhood (Td at 6 years) + Adolescent (Td at 10 years) |
| Which approach to use for vaccine introduction? |  |  |
| *Simultaneous introduction^* | *EC+C+A (S)* | *All three booster doses introduced to the target age groups in the same year* |
| Phased introduction | EC+C+A (P) | All three booster doses introduced one at a time, two years apart, to the target age groups |
| Simultaneous introduction with expanded age cohorts | EC+C+A (E) | All three booster doses introduced in the same year to the target age groups and the age groups in between (see Panel C, Figure S.2 in supplementary materials) |
| Vaccination coverage levels~ |  |  |
| All three DTPCV booster doses introduced to the target age groups in the same year: EC+C+A (Simultaneous) |  |  |
| *Default (High & Gradual)\*^* | *EC+C+A (H-G)* | *High (H) coverage (80%) & a gradual (G) increase to reach coverage (4–8 years)* |
| Conservative (Low & Gradual) | EC+C+A (L-G) | Low (L) coverage (60%) & a gradual (G) increase to reach coverage (4–8 years) |
| Optimistic (High & Accelerated) | EC+C+A (H-A) | High (H) coverage (80%) & an accelerated (A) increase to reach coverage (2–4 years) |
| Accelerated for existing age cohorts | EC+C+A (A-E) | High coverage (80%) & accelerated rollout to reach coverage for boosters that align with other vaccines provided in the existing national schedule in early childhood (measles vaccine second dose) and adolescence (HPV vaccine) |

Notes: ^Denotes the same scenario (EC+C+A with simultaneous rollout); \*Shows the inputs used as default for all the scenarios exploring number of doses and rollout approach. ~The vaccination coverage levels are estimates of what could be achieved under different operating constraints. Abbreviations: EC, Early childhood booster given at 18 months; C, Childhood booster given at 6 years; A, Adolescent booster given at 10 years; S, Simultaneous rollout approach; P, Phased rollout approach; E, Simultaneous rollout approach with expanded age cohorts in the year of introductions; H-G, high and gradual; L-G, low and gradual; H-A, high and accelerated; A-E, accelerated for existing cohorts.

### 2.4 Modelling health impact

Three primary measures of health impact are considered in the model: number of cases, number of deaths, and proportion of the population protected. Each measure is considered as a total and disaggregated by key age groups. Values are presented annually and as a total over the fifteen-year time horizon (2025–2039). The population protection is a measure of immunity, showing the proportion of the population (%) that was estimated by the model to have a protective level of antibodies by age group. For pertussis and diphtheria, immunity may be acquired through vaccination or infection [16,17]. Tetanus immunity is only vaccine derived. Both vaccine-derived and naturally acquired immunity wane over time [20]. Protection in the model is against infection for tetanus and pertussis, and against symptomatic disease for diphtheria.

All scenarios include the EC booster, which is the only DTPCV booster dose that contains pertussis (acellular or whole cell). This was chosen in collaboration with UNEPI, where priority was given to the switch from Penta to the hexavalent vaccine (DTwP–Hib–IPV– HepB)—a schedule that would include the EC booster. Therefore, model outputs for pertussis are only reported for the scenarios comparing between the baseline and one dose (EC only). For all other scenarios, model outputs focus on diphtheria and tetanus only. Because Uganda has very few reported cases and no reported outbreak of diphtheria as of 31 March 2025, the model outcome of interest for diphtheria in Uganda is the proportion of the population protected.

### 2.5 Budget impact analysis (BIA)

We conducted a BIA for each scenario, with results presented in nominal United States Dollars (USD) to facilitate international comparison and align with local reporting practices. Projected budget impact is presented annually and as a nominal total across a 10-year period (2025–2034). Results are broken down into vaccine costs, routine delivery costs, and costs of vaccine introduction activities. Budget impact projections are presented according to the source of funding as either ’full cost (donor and government)’ or ’government costs only’. Out-of-pocket costs paid by households were not included. Inflation at recent local rates was assumed for routine delivery costs but not the vaccine costs as we assume, based on trends in the previous decade and existing procurement agreements, these will remain at the agreed price negotiated by UNICEF [21]. As is standard practice, no discounting is applied to the BIA; all values are nominal [22].

The cost of vaccination is separated into 1) the incremental cost of routine vaccination delivery (by dose); and 2) the unit cost of the vaccine (by dose). The cost of routine vaccination delivery is further split by delivery location, either at a health facility or outreach site (including schools). The cost of vaccination delivery for Uganda was sourced from the Ugandan Booster Dose Costing Study (unpublished) [23]. An incremental average cost of USD 1.745 per dose delivered at a health facility and an incremental average cost of USD 1.452 per dose delivered at an outreach site was estimated (in 2025 USD). All costs are incremental to Uganda’s existing national immunisation programme. See supplementary material Tables S.2–S.3.

While the epidemiological scenarios considered a range of vaccination coverage assumptions, input from the Uganda multi-disciplinary TWG prioritised a BIA reflecting the resources required to vaccinate the total population in the target age groups. This is to support conservative planning and future funding applications. Budgeting for lower vaccination coverage levels can be scaled proportionally from the total. The BIA considers vaccination costs only and does not include the potential reduction in treatment costs due to averted cases of diphtheria, tetanus or pertussis.

### 2.6 Model fitting, uncertainty, and sensitivity analysis

Data were incorporated to fit and validate the model against observed trends, using reported cases for tetanus and GBD 2019 estimates (the most recent available at the time of this analysis) for pertussis to address underreporting. Due to the low number of reported diphtheria cases in Uganda and the role of external drivers in outbreaks, calibration focused on tetanus and pertussis only. To assess variability, the baseline model was run 1,000 times using Latin hypercube sampling across parameter ranges informed by literature and expert opinion, from which the 50 best-fitting parameter sets were selected and applied across all scenarios [24]. Model projections present 90% uncertainty intervals based on these runs, with further methodological details provided in the supplementary materials.

## 3. Results

We present our key findings on the impact of introducing different combinations of DTPCV booster doses, approaches to introduction, vaccination coverage levels, and budget impact analysis. Additional results are presented in the supplementary materials.

### 3.1 Impact of introducing the early childhood (EC) DTPCV booster only

Introducing the EC DTPCV booster in Uganda is projected to avert a total of 1,190 tetanus cases between 2025–2039, which is a 4% reduction compared to the baseline scenario (29,440 cases) (Table 2 and Figure 1). It is also projected to increase protection against tetanus and symptomatic diphtheria in the under-15-year-old population by 20% (from 25% with no DTPCV boosters to 45% with EC) and 21% (from 37% with no DTPCV booster to 58% with EC) respectively.

**Fig 1.**
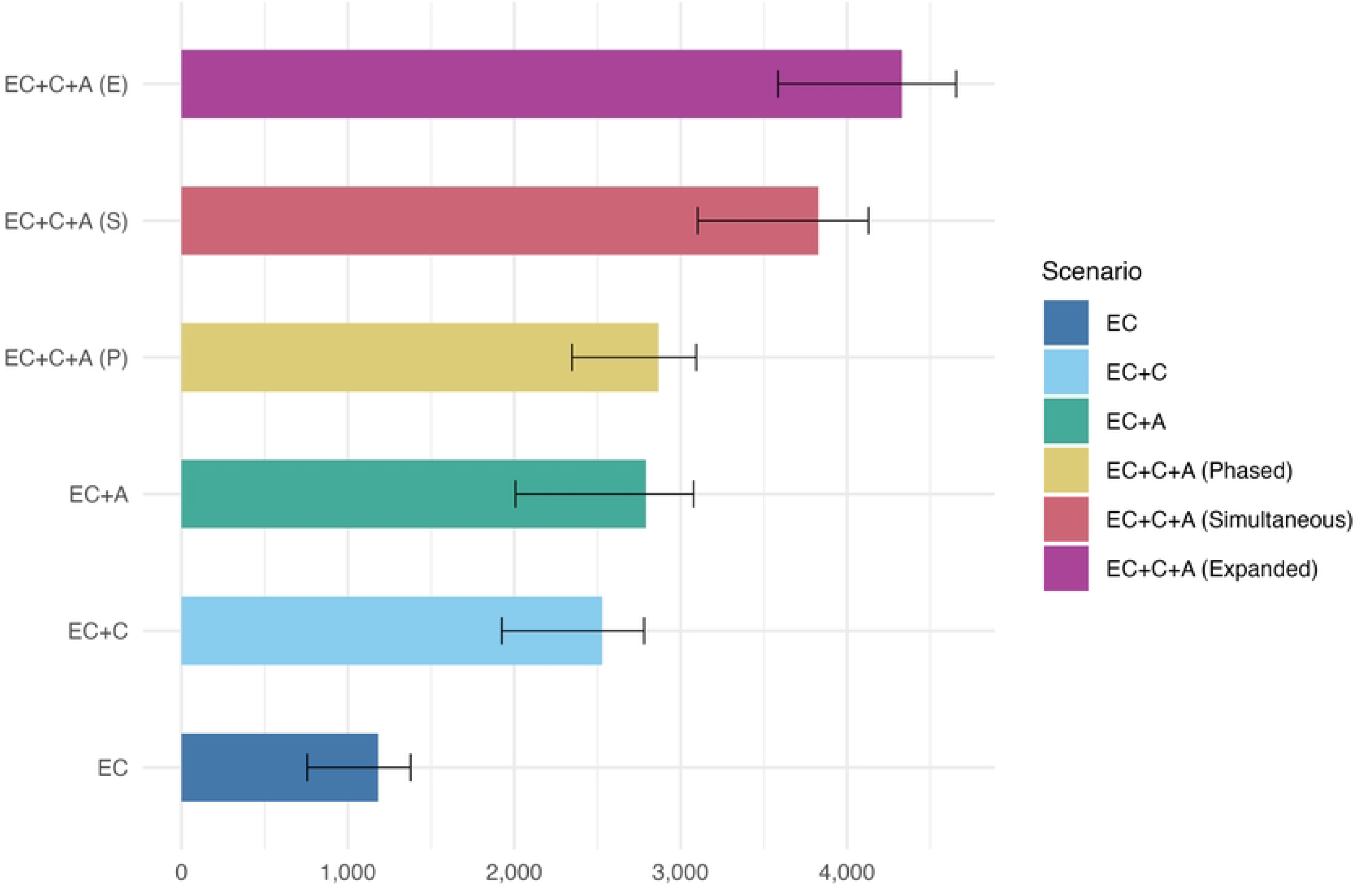
Projected total tetanus cases averted across modelled scenarios compared to baseline of no DTPCV booster doses (2025–2039) in Uganda. Figure description: Projected total number of tetanus cases averted in Uganda for each modelled scenario compared to baseline over the fifteen years from 2025 to 2039. The bar shows the median of the 50 model runs and the black whiskers on each bar display the 90% uncertainty interval. Abbreviations: EC, early childhood booster; C, childhood booster; A, adolescent booster; P, phased introduction; S, simultaneous introduction; E, expanded age cohorts in the year of introduction.

**Table 2.** Summary of impact on tetanus cases and cost to government of vaccination by type and number of DTPCV booster doses introduced in Uganda, 2025–2039*.

| DTPCV<br>Booster Doses<br>introduced | Total<br>projected<br>tetanus<br>cases<br>(2025-2039) | Average<br>annual<br>projected<br>tetanus<br>cases<br>(2025-2039) | Total<br>tetanus<br>cases<br>averted<br>compared<br>to baseline | Relative<br>multiplier<br>of tetanus<br>cases<br>averted<br>compared<br>to EC only | Incremental<br>cost (USD)<br>per<br>planned<br>dose^<br>compared<br>to baseline<br>over 10<br>years | Average<br>annual total<br>incremental<br>cost (USD) of<br>vaccination<br>compared<br>to baseline<br>(2025–2034) | Relative<br>multiplier of<br>incremental<br>cost (USD) of<br>vaccination<br>compared<br>to EC only |
| --- | --- | --- | --- | --- | --- | --- | --- |
| Baseline (no<br>booster doses) | 29,440<br>(28,890–<br>29,880) | 1,960<br>(1,930–<br>1,990) | - | - | 0.66 | - | - |
| EC only | 28,250<br>(27,680–<br>28,900) | 1,880<br>(1,850–<br>1,930) | 1,190 (760–<br>1,370) | - | 0.68 | 1,162,200 | - |
| EC+C | 26,940<br>(26,330–<br>27,650) | 1,800<br>(1,760–<br>1,840) | 2,520<br>(1,930–<br>2,780) | 2.1<br>(2.0–2.5) | 0.60 | 2,004,600 | 1.7 |
| EC+A | 26,720<br>(26,070–<br>27,540) | 1,780<br>(1,740–<br>1,840) | 2,790<br>(2,010–<br>3,070) | 2.4<br>(2.2–2.6) | 0.61 | 1,950,600 | 1.7 |
| EC+C+A (S) | 25,650<br>(25,040–<br>26,430) | 1,710<br>(1,670–<br>1,760) | 3,810<br>(3,100–<br>4,130) | 3.2<br>(3.0–4.1) | 0.58 | 2,793,000 | 2.4 |
| EC+C+A (P) | 26,580<br>(26,010–<br>27,250) | 1,770<br>(1,730–<br>1,820) | 2,860<br>(2,350–<br>3,090) | 2.4<br>(2.2–3.1) | 0.61 | 2,396,800 | 2.1 |
| EC+C+A (E) | 25,150<br>(24,530–<br>25,920) | 1,680<br>(1,640–<br>1,730) | 4,310<br>(3,590–<br>4,650) | 3.7<br>(3.4–4.7) | 0.57 | 3,364,800 | 2.9 |

Notes: Values show the median of the 50 model runs with the 90% uncertainty interval in brackets. \*Costs are shown as the total cost of vaccination (vaccines, routine delivery, and introduction activities) paid by the government, excluding donor funding. ^Cost per dose is calculated using the number of doses required for the full target population, irrespective of assumed vaccination coverage. It does not represent cost per dose actually administered. Case numbers are rounded to the nearest ten and total cost values are rounded to the nearest hundred. All costs are in nominal USD. Abbreviations: EC, early childhood booster; C, childhood booster; A, adolescent booster; S, simultaneous introduction; P, phased introduction; E, expanded age cohorts in the year of introduction.

Introducing the EC DTPCV booster in Uganda is projected to avert an estimated total of 530,400 pertussis cases between 2025–2039 compared to the baseline, which is a 9% reduction. Of these, 160,400 are estimated to be severe cases that would have sought treatment, amounting to an average of 11,000 averted hospitalised cases annually. As all other scenarios modelled include the pertussis-containing DTPCV booster in early childhood, these findings also apply to all the following scenarios.

### 3.2 Impact of introducing multiple DTPCV booster doses

Implementing all three DTPCV booster doses (EC+C+A) is projected to have the greatest impact on the proportion of the population protected against diphtheria, tetanus, and pertussis; averted tetanus cases; and averted tetanus deaths, with a lower than proportional increase in required budget, resulting in the most efficient ratio of incremental benefits to incremental costs (Table 2 and Figure 1). The baseline scenario projects a total of 29,440 tetanus cases and 15,350 deaths from 2025 to 2039 (Figure 2 and Tables S.8–S.9). This is projected to decrease by 8.5% to 26,940 total tetanus cases for EC+C; by 9.3% to 26,720 total tetanus cases for EC+A; and by 13% to 25,650 total tetanus cases for EC+C+A. Introducing all three DTPCV booster doses using a simultaneous approach is projected to avert 3.2 times the total number of tetanus cases over the fifteen years following introduction at 2.4 times the incremental cost compared to EC only (Table 2).

**Fig 2.**
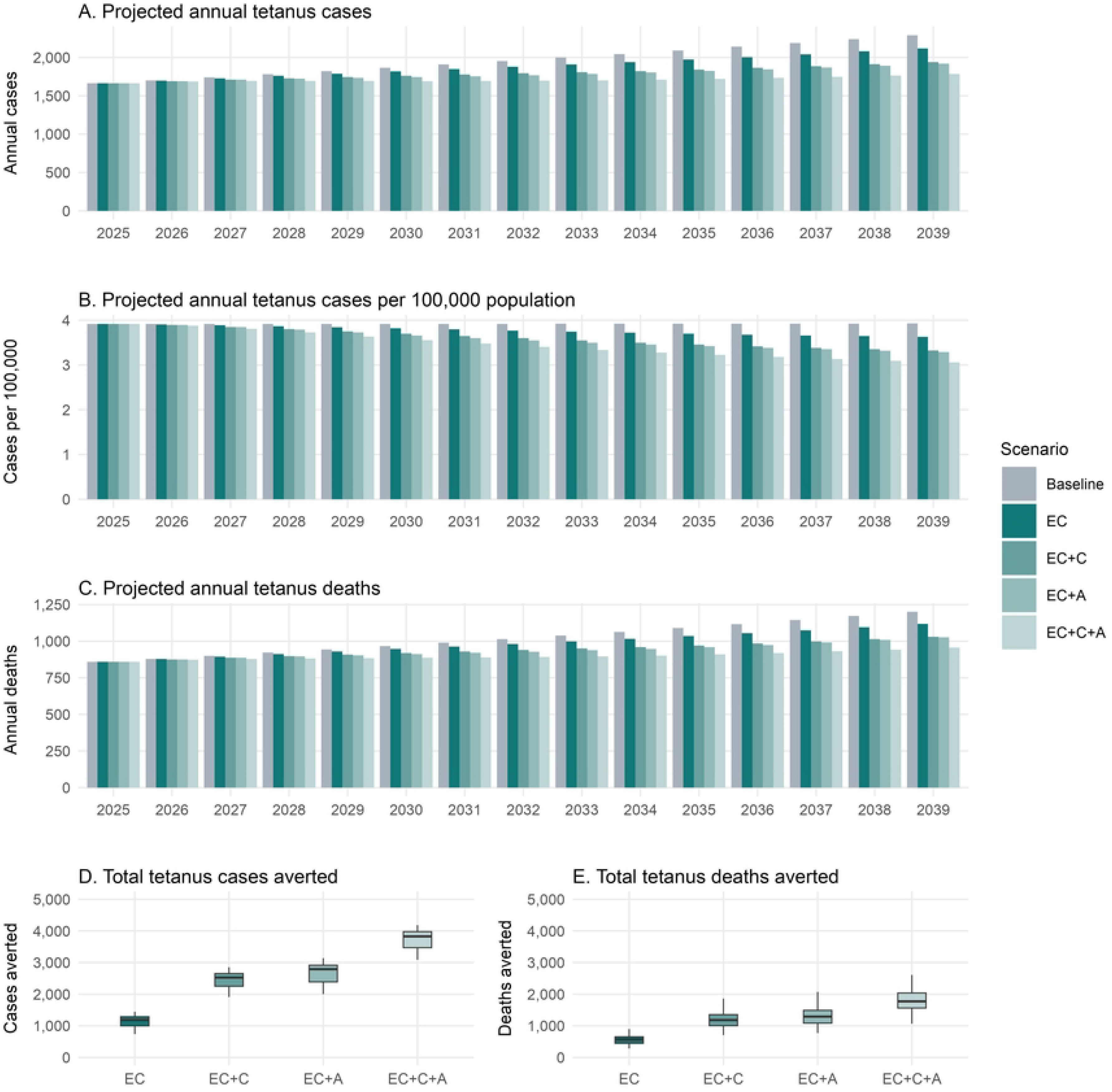
Projected annual and total cumulative tetanus cases and deaths (2025–2039) in Uganda for different combinations of DTPCV booster doses introduced simultaneously. Figure description: Panel A: Annual projected tetanus cases in Uganda from 2025 to 2039 for all ages. Panel B: Annual projected tetanus cases per 100,000 population in Uganda from 2025 to 2039 for all ages. Panel C: Annual projected tetanus deaths in Uganda from 2025 to 2039 for all ages. Panel D: Distribution of the total projected number of tetanus cases averted in Uganda from 2025 to 2039 across all model runs. Panel E: Distribution of the total projected number of tetanus deaths averted in Uganda from 2025 to 2039 across all model runs. The box limits indicate the range of the central 50% of the data, with a central line marking the median value. The whiskers show the top and bottom 25% of values. Where multiple DTPCV booster doses are introduced, this is done simultaneously. Abbreviations: EC, early childhood booster; C, childhood booster; A, adolescent booster; P, phased introduction; S, simultaneous introduction; E, expanded age cohorts in the year of introduction.

Simultaneous introduction of all three DTPCV booster doses is projected to increase population protection against symptomatic diphtheria by 2.2 times and against tetanus infection by 2.8 times in under 15-year-olds compared to no DTPCV booster doses, after 15 years of delivery (in 2039) (Figures S.8–S.10). Figure 3 shows the full breakdown of the projected population protection for tetanus by age group from 2025 to 2039. The baseline scenario projects that 25% of under 15-year-olds in Uganda will be protected against tetanus in 2039. Following the introduction of DTPCV booster doses, the proportion of under 15-year-olds in Uganda projected to be protected against tetanus infection increases to 45% for EC only; to 62% for EC+C; to 56% for EC+A; and to 69% for all three doses (EC+C+A). The baseline scenario projects that 37% of under 15-year-olds in Uganda will be protected from symptomatic diphtheria (Figures S.9–S.10). This is projected to increase to 58% for EC only; to 77% for EC+C; to 70% for EC+A; and to 81% for all three doses (EC+C+A). Compared to EC only, introducing all three DTPCV boosters is projected to result in 1.4 and 1.5 times the population protected against symptomatic diphtheria and tetanus infection respectively, by 2039.

**Fig 3.**
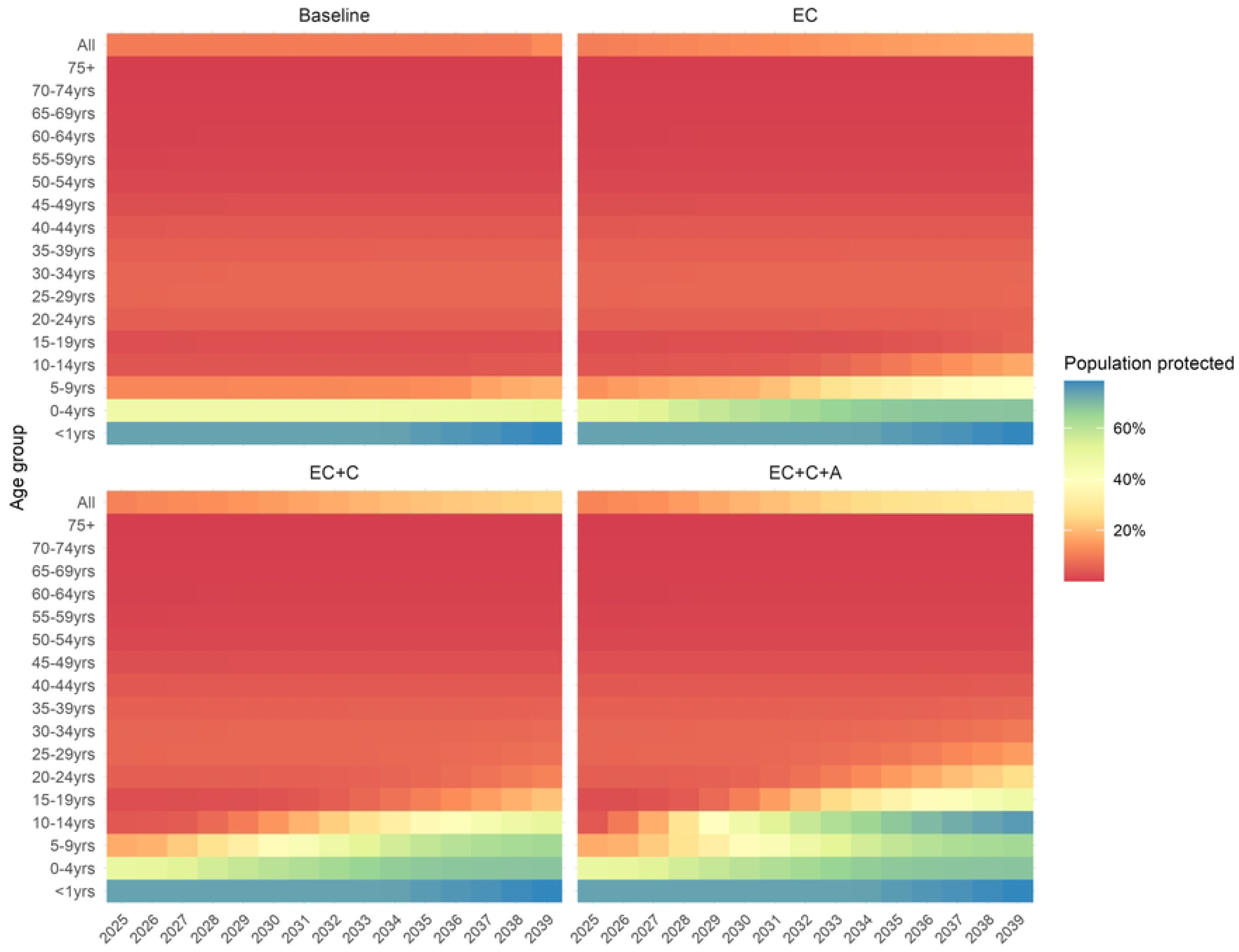
Projected proportion (%) of the Ugandan population protected against tetanus, by age group, for different target age groups and number of DTPCV booster doses. Figure description: Age groups are presented in 5-year bands, with additional categories shown for All ages (top row of the heatmap) and children under one year of age. The latter category is a subset of the age group for children under five years of age. The panels show different combinations of the number and target-age groups for DTPCV booster doses: Baseline, EC only, EC+C, and EC+C+A (Table 2). Shading shows the percentage of the Ugandan population protected against tetanus, from low (red) to high (blue). Abbreviations: EC, early childhood booster; C, childhood booster; A, adolescent booster; P, phased introduction; S, simultaneous introduction; E, expanded age cohorts in the year of introduction.

The impact of expanding the target age cohorts in the year of DTPCV booster dose introduction (simultaneous introduction with expanded age cohorts) depends on the coverage that can be attained. Assuming 35% vaccination coverage in the year of introduction for all age cohorts, a one-time expansion of the age cohorts is projected to avert 500 more tetanus cases than the simultaneous approach alone over the fifteen years following introduction. With finite resources for introducing new vaccines, the model findings suggest that either prioritising investment for achieving high coverage sooner for the target age groups or expanding the age cohorts in the year of introduction will have similar effects, depending on what coverage can be achieved for these targeted and expanded age cohorts.

### 3.3 Impact of different approaches to vaccine introduction

All three approaches to DTPCV booster dose introduction (Table 1, Figure S.11) are estimated to result in a substantial decrease in tetanus cases and deaths compared to the baseline scenario of not introducing any DTPCV boosters. However, phasing the introduction of each DTPCV booster dose is a missed opportunity to increase population protection and is projected to take 14 years longer to reach the same level of population protection as a simultaneous introduction of all DTPCV booster doses (Figure 4, Figure S.12). The simultaneous introduction approach is projected to result in 30% more tetanus cases averted between 2025 and 2039 than the phased approach, at 15% higher cost. The other potential benefits of the simultaneous approach include improved logistical efficiencies and economies of scale, which could reduce the costs of introduction activities.

**Fig 4.**
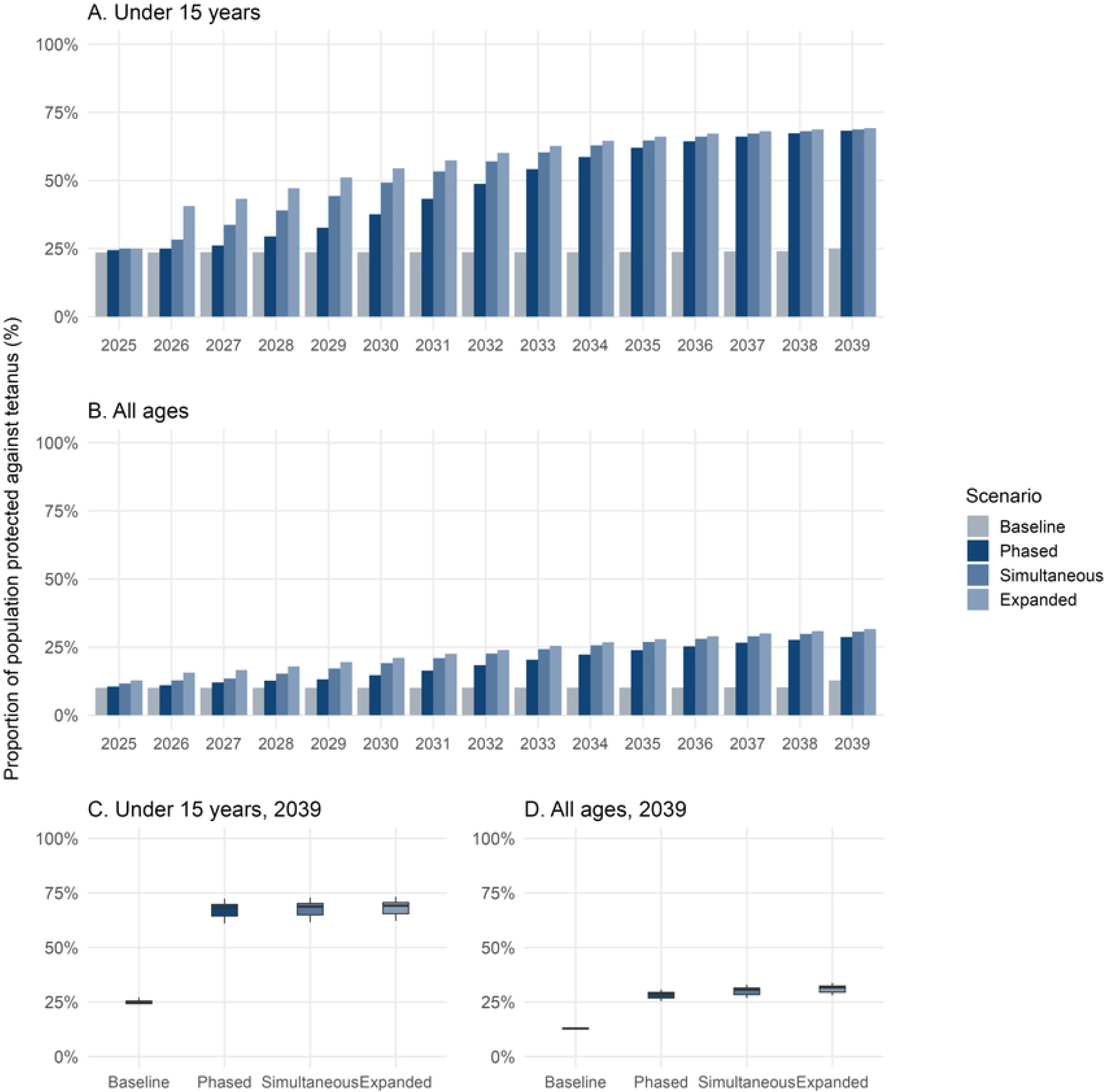
Projected proportion (%) of the Ugandan population protected against tetanus, for different age groups, fifteen years following introduction of three DTPCV booster doses using different approaches to introduction. Figure description: Panel A: Proportion of the population under 15-years of age in Uganda that is projected to be protected against tetanus by scenario over time. Panel B: Proportion of the population across all age groups that is projected to be protected against tetanus by scenario over time. Panels C and D show the distribution of the values for the projected population protection in Uganda in 2039 for those under 15 years of age and all ages respectively. The box limits indicate the range of the central 50% of the data, with a central line marking the median value. The whiskers show the top and bottom 25% of values across all model runs. Abbreviations: EC, early childhood booster; C, childhood booster; A, adolescent booster; P, phased introduction; S, simultaneous introduction; E, expanded age cohorts in the year of introduction.

### 3.4 Impact of variations in vaccination coverage

For all introduction scenarios, the expected vaccination coverage and years to reach target vaccination coverage are important assumptions that substantially impact the potential benefits of introducing DTPCV booster doses. The optimistic vaccination coverage scenario averts approximately 50% more cases of tetanus over the fifteen years following DTPCV booster dose introduction than the conservative scenario (Table S.10, Figure S.15).

### 3.5 Budget impact

As expected, the more DTPCV booster doses introduced, the higher the projected total cost of vaccination (Figure 5). However, the increases are not linear as the introduction costs are spread over multiple doses and the mix of delivery location and costs vary by DTPCV booster dose. The projected cost to the Government of Uganda over ten years for the introduction of a single DTPCV booster dose (early childhood) is USD 11.6 million (Table 3), of which USD 3.4 million is the cost of the vaccine only (no delivery costs). The projected total cost of adding a second DTPCV booster dose (either childhood or adolescent booster) is 1.7 times the projected cost of introducing only the early childhood DTPCV booster (USD 19.5–20 million total; USD 5–5.2 million for vaccine costs only). The projected cost of introducing all three DTPCV boosters is two to three times the cost of introducing the early childhood booster only, depending on the rollout approach taken (USD 23.9–33.6 million total; USD 6.8 million for vaccine costs only). The projected cost of introducing all three DTPCV booster doses in a simultaneous approach is 2.4 times that of introducing the early childhood DTPCV booster only (USD 27.9 million; USD 6.8 million).

**Fig 5.**
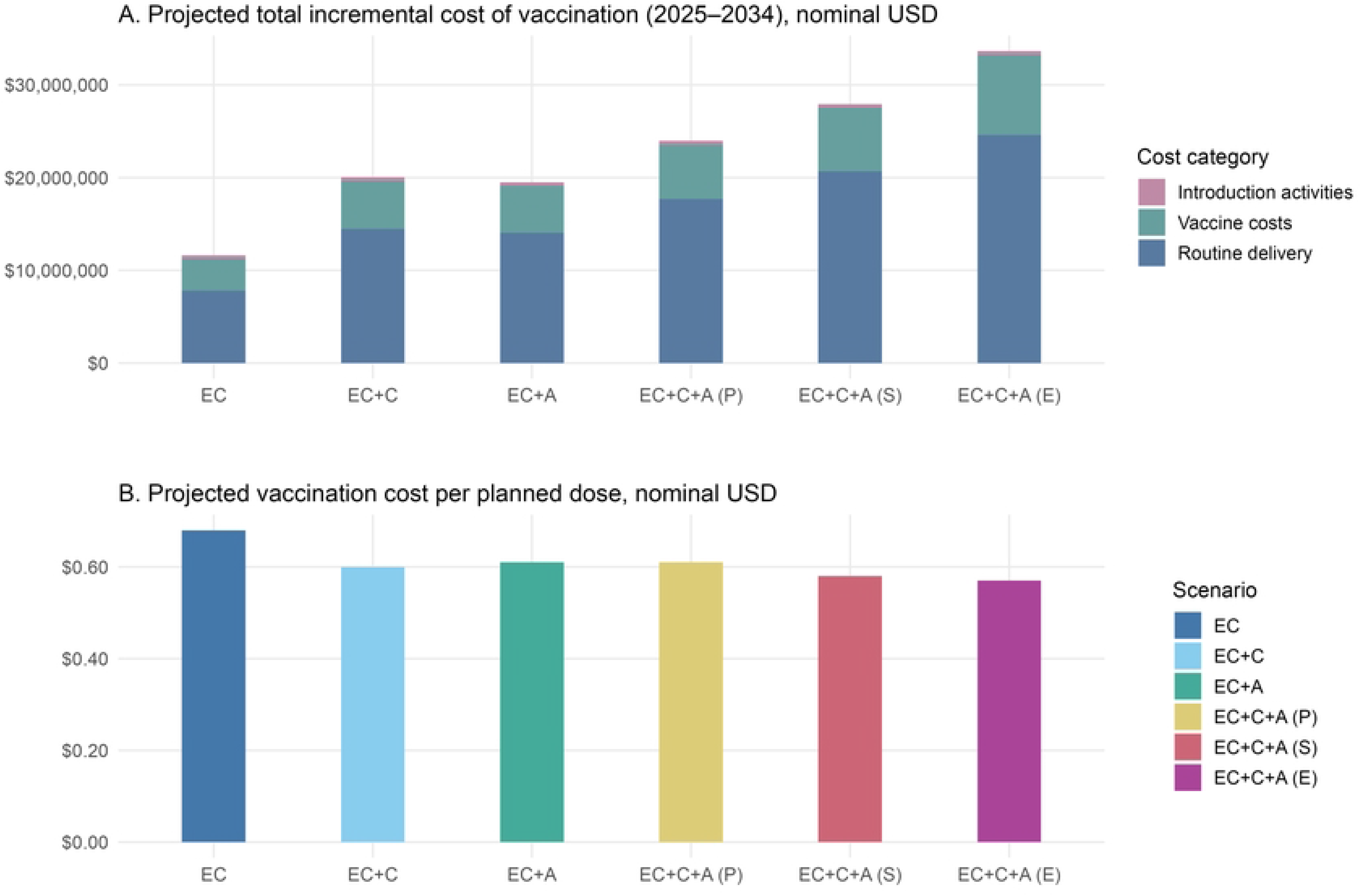
Projected incremental cost of DTPCV booster dose vaccination in Uganda, total and per planned dose (government only), compared to baseline over ten years (2025– 2034) Figure description: Panel A shows the projected total incremental government cost of vaccination by scenario and cost category over 2025–2034. Panel B shows the projected government vaccination cost per planned dose by scenario. Cost per planned dose is calculated using the number of doses required for the full target population, irrespective of assumed vaccination coverage, and does not represent the cost per dose administered. Abbreviations: EC, early childhood booster; C, childhood booster; A, adolescent booster; P, phased introduction; S, simultaneous introduction; E, expanded age cohorts in the year of introduction.

**Table 3.** Projected incremental cost (nominal USD) per planned DTPCV booster dose (A) and total incremental nominal cost (B) of different vaccination strategies compared to baseline (primary series only) over ten years (2025–2034) in Uganda.

| Scenario | Government only |  | All payers<br>(government and donor) |  |
| --- | --- | --- | --- | --- |
|  | Vaccines only<br>(Nominal USD) | Total^<br>(Nominal USD) | Vaccines only<br>(Nominal USD) | Total^<br>(Nominal USD) |
| (A) Projected incremental cost per planned dose |  |  |  |  |
| Baseline* | 0.20 | 0.66 | 0.80 | 2.88 |
| EC only | 0.20 | 0.68 | 0.80 | 3.00 |
| EC+C (S) | 0.16 | 0.60 | 0.46 | 2.50 |
| EC+A (S) | 0.16 | 0.61 | 0.48 | 2.53 |
| EC+C+A (S) | 0.14 | 0.58 | 0.35 | 2.34 |
| EC+C+A (P) | 0.15 | 0.61 | 0.41 | 2.49 |
| EC+C+A (E) | 0.15 | 0.57 | 0.39 | 2.30 |
| (B) Projected total incremental nominal cost (2025–2034) |  |  |  |  |
| Baseline* | 10,337,600 | 34,030,900 | 41,350,400 | 149,047,600 |
| EC only | 3,409,600 | 11,621,900 | 13,639,000 | 51,146,800 |
| EC+C (S) | 5,185,600 | 20,046,100 | 15,414,900 | 83,141,400 |
| EC+A (S) | 5,067,300 | 19,505,500 | 15,296,600 | 81,104,000 |
| EC+C+A (S) | 6,842,900 | 27,929,500 | 17,072,500 | 113,098,800 |
| EC+C+A (P) | 5,886,200 | 23,967,500 | 16,115,800 | 98,482,200 |
| EC+C+A (E) | 8,669,100 | 33,647,700 | 22,884,300 | 136,601,200 |

Notes: All costs are in nominal USD. Costs per planned dose are calculated using the number of doses required for the full target population, irrespective of assumed vaccination coverage. \*Baseline values (cost of primary series only) shown for reference. ^Total shows the ten-year nominal total, including vaccine costs, routine delivery costs, and cost of introduction activities. Abbreviations: EC, early childhood booster; C, childhood booster; A, adolescent booster; S, simultaneous introduction; P, phased introduction; E, expanded age cohorts in the year of introduction.

Economies of scale are seen when comparing the cost per DTPCV booster dose across different introduction approaches and numbers of doses (Table 3). The early childhood DTPCV booster is projected to cost USD 0.68 per planned dose. The cost per planned dose when introducing two DTPCV booster doses (either EC+C or EC+A) is projected to be USD 0.60 and USD 0.61, respectively. Introducing all three DTPCV booster doses simultaneously is projected to cost USD 0.58 per planned dose. While the phased approach saves on total cost and spreads the burden of budget increases over six years, it leads to a higher cost per planned dose (USD 0.61) and reduced health impacts. The introduction of three DTPCV booster doses with expansion to additional age cohorts in the year of introduction has a substantial cost in the first year and the highest projected cost overall (USD 33.6 million over ten years) (Figure 6). However, its cost per planned dose is projected to be the lowest among the introduction scenarios (USD 0.57). The complete projected budget impact by payer, cost category, and year is provided in the supplementary materials (Table S.12). All costs are based on the total population in target age groups and not the actual coverage attained, thus these costs represent the estimated upper bound. If the procurement strategy is to purchase fewer vaccines or rollover additional vaccine supply from previous years, actual costs would be lower.

**Fig 6.**
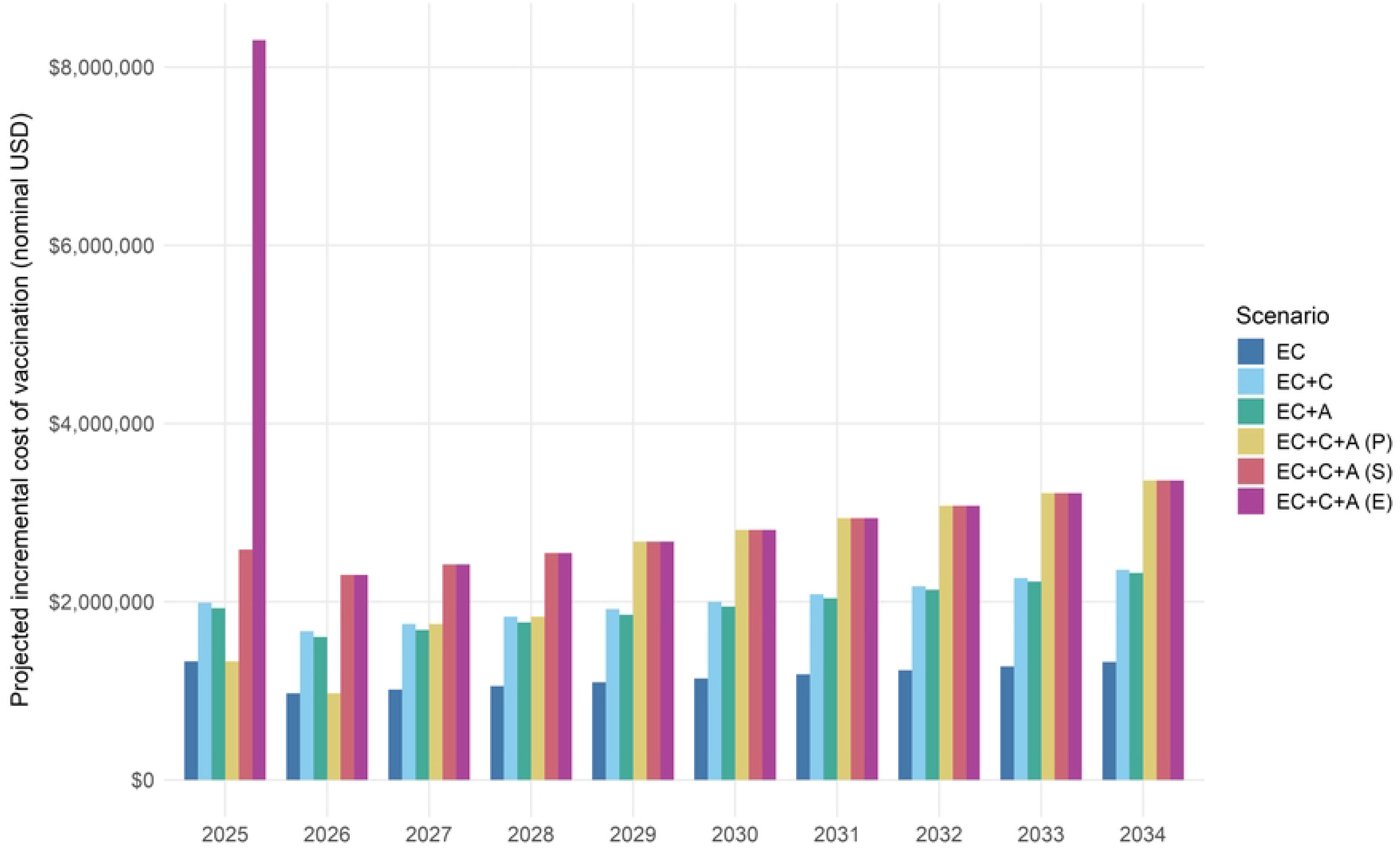
Projected annual incremental government cost of DTPCV booster dose vaccination by scenario compared to baseline in Uganda, 2025–2034. Figure description: Bars show projected annual incremental vaccination costs paid by the Government of Uganda compared with baseline. Abbreviations: EC, early childhood booster; C, childhood booster; A, adolescent booster; P, phased introduction; S, simultaneous introduction; E, expanded age cohorts in the year of introduction.

## 4. Discussion

Diphtheria, tetanus, and pertussis remain diseases of public health importance though few African countries have childhood vaccination schedules that optimise protection into adulthood. We built an integrated multi-disease epidemiological-economic model for diphtheria, tetanus, and pertussis to explore the health and budget impact of introducing DTPCV booster doses and applied it for this first-ever use case in Uganda. We considered country-defined scenarios about different number and combination of DTPCV booster doses, introduction approaches, and the implications of vaccination coverage on outcomes in Uganda. Introducing all three DTPCV booster doses (EC+C+A) is projected to have the greatest impact on the proportion of the Ugandan population protected against diphtheria, tetanus, and pertussis; averted tetanus cases; and averted tetanus deaths, without a correspondingly large increase cost compared to introducing EC only. The cost per planned dose is USD 0.58 for EC+C+A compared to USD 0.68 for EC only, with an average annual total incremental cost from the government perspective of USD 2.79 million and USD 1.16 million respectively. Simultaneous introduction of all three DTPCV booster doses is projected to avert approximately 30% more tetanus cases at only 15% higher cost than phased introduction. These findings suggest that simultaneously introducing all three DTPCV booster doses has the greatest health impact and is the best value-for-money option under the modelled assumptions. If only two DTPCV booster doses are affordable or feasible (i.e. Penta4/Hexa4 + 1 dose of Td), it is important to consider local priorities, epidemiological risk groups, and operational feasibility when deciding which doses to introduce. While there is little difference in the proportion of the population protected between EC+C versus EC+A across the whole population, EC+C is predicted to be more effective than EC+A in providing protection against diphtheria and tetanus in under 15-year-olds. Introducing only the EC dose still provides notable health benefits over the primary series, with the most modest budget impact of the scenarios considered in our analysis.

Introducing the recommended three DTPCV booster doses is projected to approximately double population protection against symptomatic diphtheria and triple protection against tetanus in under 15-year-olds in Uganda ten years post-introduction—important findings given Uganda’s high tetanus burden and the diphtheria resurgence across Africa since 2023 [13]. However, measuring and prioritising health impact differs meaningfully across these three diseases. For tetanus, where case numbers are low, demonstrating the added value of interventions through cases or DALYs averted is inherently difficult. For diphtheria, where endemic transmission is absent, the benefit of vaccination lies in increased population protection against future outbreaks, which is a benefit that, while not immediately realised, is essential for reducing outbreak-associated cases and deaths. Yet quantifying the relationship between population immunity and outbreak risk remains methodologically complex, as does assigning an economic value to incremental gains in protection. Unlike DALYs averted, for which decision thresholds are well established, no widely accepted framework exists for valuing immunity as an outcome measure in economic evaluation. Future research should explore methodologies for quantifying this benefit and estimating the return on investment of preventing or attenuating future outbreaks.

Research emphasises that reaching and maintaining high vaccination coverage is crucial for achieving health impact and preventing outbreaks [4,25]. For all our scenarios, the expected vaccination coverage level and years to reach this coverage level are important assumptions affecting model outputs. The optimistic scenario averts approximately 50% more cases of tetanus over the fifteen years following introduction than the conservative scenario. Several studies have been conducted in Uganda on the barriers to uptake of vaccines [26,27]. Health system barriers raised by these studies were mostly logistical in nature and reflect inadequate resourcing of immunisation efforts. In our scenarios, we considered the role of coverage on the impact that can be attained through the introduction of DTPCV booster doses. A further analysis that would be valuable to explore is a comparison of the feasibility, cost, and cost-effectiveness of increasing coverage of diphtheria-tetanus-pertussis vaccine doses that are already part of the routine immunisation schedule versus introducing new doses at existing (or potentially lower) coverage levels. With an increasing portfolio of available vaccine doses to choose from and prioritise, an examination of such trade-offs could support decision-making in resource-constrained settings and demonstrate the value of attaining high coverage.

From the Government of Uganda (payer) perspective, the average annual incremental cost over 10 years of routine delivery of three DTPCV booster doses, introduced simultaneously is USD 2.79 million. Of this, USD 1.16 million (41.6%) is for the EC booster, which is the most expensive vaccine of the three to purchase but also offers protection against hepatitis B and *Haemophilus influenzae* type b in addition to diphtheria, tetanus, and pertussis. The projected financial cost of vaccination per planned DTPCV dose is USD 0.58 for all three introduced simultaneously compared to USD 0.68 for EC only. Therefore, although the total budget required is higher for all three boosters, introduction of the three boosters offers better value per planned dose than introducing EC only. These values per dose represent the amount projected to be paid by the Ugandan government, excluding co-financing mechanisms. If Uganda’s GNI growth remains consistent, the country is unlikely to transition out of the most generous phase of donor support from Gavi (initial self-financing status) during the ten-year period of the budget impact analysis conducted in this study. However, given the precarious nature of the current global health funding landscape, understanding the full cost of new interventions is important for assessing the sustainability should circumstances change. The WHO’s Global Health Expenditure Database reports that Uganda’s current health expenditure (CHE) was USD 2,200 million in 2023 [28]. The domestic general government health expenditure (GGHE-D) is 22% of CHE. Uganda’s CHE per capita is USD 45 and GGHE-D per capita is USD 10 [28]. The large contribution of external funding to the total health budget presents challenges to sustainability and prioritisation in the medium- and long-term. If there were no donor contributions or co-financing mechanisms in place, our analysis projects the annual average cost to the Ugandan government over the first ten years following introduction could increase to USD 5.1 million and USD 11.3 million for the EC booster only and all three DTPCV doses, respectively, depending on the approach to demand calculation.

Implementation of the Ugandan National Immunisation Strategy (NIS) 2024–2028 is estimated to cost a total of USD 1,484 million (nominal) over five years of the strategy, averaging USD 296.8 million annually [29]; this is 14% of Uganda’s CHE. The three-dose primary series of pentavalent vaccine alone is projected to cost 1.5% of the total annual EPI budget (USD 4.37 million per year) over the next five years. All three DTPCV booster doses, introduced simultaneously, are projected by our analysis to comprise 2.3–3.8% of the total EPI resource requirement, depending on the approach to demand estimation. The limited resources for health and competing priorities for these, lead to the desire to understand the health benefits of introducing some, but not all DTPCV booster doses. If only two DTPCV booster doses are affordable, this should include the early childhood pertussis-containing dose with the choice between the childhood (C) or adolescent booster (A) doses informed by factors such as prioritisation of at-risk populations, operational feasibility, and synergies with other vaccines in the existing national immunisation schedule. For example, if the highest disease prevention priority is tetanus (especially maternal and neonatal tetanus), EC+A is projected to provide protection longer into adulthood than EC+C. If local data suggests under 15-year-olds as a high-risk group (e.g. risk of severe diphtheria in children, high risk of tetanus related to male circumcision), EC+C is projected to provide greater protection in these ages than EC+A.

When using the findings from this work for decision-making it is important that they are considered alongside other data, literature, local priorities, current realities, and expert knowledge. The collaborative development of scenarios and key inputs with UNEPI and local experts was prioritised to reflect the local context and demonstrate the potential impact of a range of operational delivery factors such as target vaccination coverage, service delivery location, and number of years to reach target vaccination coverage. However, the actual rollout of any DTPCV booster doses may differ from the assumed vaccination coverage and delivery strategies modelled. Logistics, risk profiles, and behavioural factors should also be considered when interpreting projections for decision-making. For example, a change in the proportional split in delivery locations would affect the total cost, as outreach delivery sites are estimated to be cheaper per dose delivered than health facility-based delivery [23]. If a new DTPCV booster is introduced alongside an existing vaccine (for example the adolescent DTPCV booster may be added at the same age-based vaccination schedule point as human papillomavirus vaccine), there could be further operational efficiencies and cost reductions than those presented. The findings from this study were used to inform UNITAG’s 2025 New Vaccines Prioritisation, contributing to decision-making on the introduction of DTPCV booster doses in Uganda [30]. The UNITAG recommended all three DTPCV booster doses be introduced.

### 4.1 Limitations

Limitations of the underlying disease models are discussed in the respective publications [16,17,20]. The application of the integrated epidemiological-economic model in Uganda comes with some limitations. First, model reliability is constrained by data availability and quality. For tetanus, calibration was based on Ugandan WHO/UNICEF Joint Reporting Form (JRF) data, which, despite generally high reporting rates for this severe disease, remain subject to errors including overreporting, misclassification, and underreporting as documented in field investigations [10]. For pertussis, the absence of reliable national surveillance, serological, or incidence data necessitated calibration to GBD estimates and expected epidemic patterns. However, pertussis burden is inherently difficult to measure due to underreporting, misclassification, and asymptomatic infection, meaning GBD estimates may embed systematic uncertainties. In the absence of Uganda-specific data, we balanced GBD estimates with plausible disease dynamics and historical vaccination coverage. Limited national surveillance and serological data for diphtheria precluded direct model calibration; instead, the model was validated against settings with available surveillance data and adapted using Uganda-specific demographics and immunisation inputs. These limitations introduce uncertainty, and results should therefore be interpreted cautiously, with analyses framed as scenario exploration rather than prediction.

Second, the integrated disease model was applied nationally. While a national perspective provides robust population-level projections, it does not capture subnational variation. There may be important differences in characteristics such as vaccination coverage, population contact patterns, disease exposure, treatment-seeking behaviour, and treatment availability. Third, the timeframe for the epidemiological model is fifteen years, and the amount of uncertainty in model estimates increases the further into the future projections are made. However, vaccination offers benefits over a longer period than the fifteen-year timeline modelled. Therefore, our findings show a conservative estimate of what health benefits may be attained in the next fifteen years rather than the lifetime benefits of DTPCV booster doses. Fourth, the projected budget impact is for the total population in the target age groups. However, this is likely to be an overestimate if the procurement policy or approach to demand estimation is based on actual vaccine doses to be delivered or if there is stock rollover between years to minimise wastage [22,31]. Finally, our unit cost for vaccine price of the EC booster assumed that the pentavalent vaccine would be used. If a DTP formulation without the additional antigens was used instead, the cost would be about a quarter of the pentavalent price. While this would not substantially affect the cost estimates from the government perspective (as donor co-financing would reduce the price per dose of pentavalent vaccine to a similar amount), the total resources required would be much lower.

### 4.2 Conclusion

The introduction of DTPCV booster doses in Uganda is projected to substantially increase population immunity and reduce disease burden, with simultaneous delivery of all three doses providing the greatest health gains and lowest cost per dose under our modelled assumptions. Our findings provide timely, country-specific evidence to inform national immunisation policy, supporting the prioritisation of comprehensive booster schedules over incremental approaches. The integrated modelling framework used here offers a practical and adaptable tool to support evidence-based vaccine policy across similar settings. As countries expand their immunisation programmes amid constrained resources and evolving disease threats, such approaches can help guide strategic investments that maximise long-term health impact and system resilience.

## Author contributions

R.A. Hounsell: conceptualisation, data curation, formal analysis, investigation, methodology, software, validation, visualisation, writing – original draft. S.P. Silal: conceptualisation, data curation, formal analysis, investigation, methodology, software, validation, writing – review & editing. J. Norman: conceptualisation, data curation, formal analysis, investigation, methodology, software, validation, visualisation, writing – review & editing. R. Monyake: formal analysis, investigation, methodology, software, visualisation, writing – review & editing. A. Immaculate: conceptualisation, data curation, formal analysis, investigation, validation, writing – review & editing. A. Besigye: data curation, formal analysis, investigation, validation, writing – review & editing. G. Biroma: data curation, formal analysis, investigation, validation, writing – review & editing. J. Magoola: data curation, formal analysis, investigation, project administration, validation, writing – review & editing. N. Kishore: conceptualisation, data curation, formal analysis, investigation, methodology, project administration, validation, writing – review & editing. S.E. Pallas: conceptualisation, data curation, investigation, methodology, validation, writing – review & editing. K. Ward: conceptualisation, data curation, formal analysis, investigation, funding acquisition, methodology, project administration, validation, writing – original draft.

## Acknowledgements

We would like to extend our thanks to the Technical Expert Group who served as scientific advisors and provided input on model development, epidemiology and immunology of disease, application features and dissemination: Dr Ampeire Immaculate, Ugandan National Immunization Program, Ministry of Health, Government of Uganda; Dr Anna Acosta, Meningitis and Vaccine Preventable Diseases Branch at U.S. Centers for Disease Control and Prevention (CDC); Dr Annet Kisakye, Expanded Program on Immunization, World Health Organization, Uganda; Prof Paula Mendes Luz, Instituto Nacional de Infectologia Evandro Chagas (INI); Dr Todi Mengistu, Measurement, Evaluation and Learning (MEL) team at Gavi, the Vaccine Alliance; Dr Helen Quinn, National Centre for Immunisation Research and Surveillance (NCIRS); University of Sydney Children’s Hospital Westmead Clinical School; So Yoon Sim, Value of Vaccines, Modeling & Economics (VoV) team, Immunization Analysis & Insights (IAI) unit, Department of Immunization, Vaccines and Biologicals (IVB), World Health Organization; Dr Rania Tohme, Hepatitis B and Tetanus Team in the Global Immunization Division at the U.S. Centers for Disease Control and Prevention (CDC); and Prof Rudzani Muloiwa, Vaccines for Africa Initiative (VACFA), Faculty of Health Sciences, University of Cape Town, South Africa. We appreciate the unwavering support of the Government of Uganda National Expanded Programme on Immunization (UNEPI) leadership throughout the entirety of this work.

## Disclaimer

The findings, conclusions, and views expressed in this article are those of the authors and do not necessarily represent the official position of the US Centers for Disease Control and Prevention (CDC), the US Department of Health and Human Services, the African Field Epidemiology Network (AFENET), or the Republic of Uganda Ministry of Health

## Data availability

All data underlying the findings reported in this study are available in the manuscript, Supporting Information, or publicly accessible repositories. Model input parameters and assumptions used for the Uganda analyses are provided in S1 Appendix. Publicly available epidemiological, demographic, vaccination, and economic input data are identified with their sources in the manuscript and S1 Appendix.

Use of artificial intelligence tools. During manuscript preparation, the authors used ChatGPT Edu (OpenAI), accessed through the University of Oxford’s institutional ChatGPT Edu workspace, to assist with editorial review and language revision. All AI-assisted suggestions were critically reviewed and revised by the authors. No generative AI was used to generate or analyse study data, perform the modelling analyses, or generate scientific conclusions. The authors take full responsibility for the final manuscript.

## Supporting information

S1 Appendix. Supplementary materials on methodology and results. Supplementary materials on scenarios, inputs, and assumptions; model fitting, uncertainty, and sensitivity analysis; supplementary findings from the epidemiological analysis; supplementation findings from the budget impact analysis.

